# What are the experiences, perceptions and training needs of inpatient general nursing staff on Continuous Glucose Monitoring?

**DOI:** 10.64898/2026.10.14.26365417

**Authors:** Busra Donat Ergin, Katharina Mattishent, Deborah Holder, Anne-Marie Minihane, Bethany Kelly, Richard Holt, Helen R. Murphy, Ketan Dhatariya, Michael Hornberger

## Abstract

**Background:** Although point-of-care testing is the standard method for inpatient glucose monitoring; it is associated with patient discomfort and late detection of dysglycaemic events. Continuous glucose monitoring (CGM) may improve glycaemic management; however, limited staff education and experience among may hinder its implementation.

**Aim:** To investigate experience, perceptions and training needs on CGM use among inpatient nursing staff across diverse hospital wards.

**Methods:** This is a single-centre, 38-item online survey recruited inpatient nurses in a hospital in the UK. Participants were invited via the e-newsletter and posters; the survey remained open for five months.

**Results:** Nineteen nurses reported positive perceptions of inpatient CGM. Most considered CGM reliable (77.8%) and preferred CGM combined with POCT (66.7%). Views on its impact on workload and patient-room entries were mixed (30% agreed, 30% disagreed). Training needs included sensor placement (92.9%) and alarm management (78.6%).

**Conclusion:** Nurses viewed CGM favourably but highlighted important training needs for inpatient use.

**Key points:**

1. Inpatient nurses generally viewed CGM positively and considered it as a reliable tool.
2. Most nurses preferred using CGM alongside POCT, while views on its impact on patient-room entries were mixed.
3. Most nurses identified further training needs, particularly for sensor placement and CGM alarm management.
4. The small, single-site voluntary sample limits generalisability; findings should be confirmed in larger, multicentre studies.

## Introduction

Diabetes and stress-related hyperglycaemia are common in hospital inpatients with diabetes being present in 20-34% of hospitalised adults in general medicine and surgery units [1], [2]. Dysglycaemia and high glucose variability are associated with longer hospital stays, chronic complications of diabetes and mortality [1], [3], [4], requiring, therefore, close management in hospital settings.

Bedside capillary point of care testing (POCT) is the standard and usual glucose management tool for inpatients with diabetes [2], [5]. Although it is still a gold standard for inpatient care; some practical, patient-centred and nursing-related challenges remain. Documentation burden for staff, limited cooperation in particular patient groups (e.g. paediatrics, mental health conditions, cognitive impairment) and patient discomfort due to repeated bedside testing have been listed as some of the limitations of POCT in hospital settings [6], [7], [8].

Some of these challenges may be overcome through the use of technology [9] such as continuous glucose monitoring (CGM) which may help glucose management in inpatients by allowing more granular dysglycaemia monitoring whilst reducing POCT discomfort [9], [10]. Previous research shows evidence of improved glycaemic control and reduced hypoglycaemia with CGM use in addition to POCT, compared to POCT alone [5], [10]. Despite the significant benefits of CGM, there are also considerable barriers to its widespread use in inpatients, which include: i) costs for the healthcare system; ii) lack of regulatory clearance for inpatient use; iii) alarm/alert fatigue among patients and staff; iv) lack of clinical practice guidelines; and v) lack of education and limited training opportunities on CGM use among healthcare professionals [11], [12], [13].

Indeed, previous research identified inexperience and lack of training of healthcare professionals as one key barrier for CGM use in hospital settings [12], [14], [15]. Previous survey studies in intensive care unit (ICU) nurses [16] and inpatient diabetes specialist nurses (DSN) [17] have consistently reported positive perceptions and acceptance of CGM use in the inpatient setting. ICU nurses also reported improved workflow in intensive care units with CGM and preferred its use over POCT alone for inpatient glucose monitoring [16]. However, to our knowledge, there are no studies investigating the experience, perceptions and training needs for CGM of general, i.e. non-DSN or ICU, nursing staff. General inpatient nursing staff increasingly come across CGM and yet it is not clear what their experience, perceptions and training needs for this technology are.

The current study attempts to fill this gap by conducting a single-centre general nursing survey in a tertiary hospital in the UK. The study aimed to investigate inpatient nurses’ experience, perceptions, and training needs regarding CGM use, while also exploring their views on traditional POCT. Considering the substantial potential benefits of CGM use, including reduced staff workload, improved patient comfort through fewer POCT measurements, and early detection of hypoglycaemia [16], [18], understanding its implementation in routine clinical practice is of clear importance. We hypothesised that general nursing staff would have some experience with CGM sensors within their routine clinical practice, while relying mostly on POCT as standard of care. We also hypothesised that general ward staff would have limited knowledge of CGM use compared with POCT and would have significant training needs.

## Methodology

### Participants and setting

The participant group consisted of nurses or student nurses over 18 years working at the Norfolk and Norwich University Hospital (NNUH), United Kingdom. The participants needed to be fluent in written and spoken English to join the study. Nurses or student nurses from all clinical units including intensive care units (ICU) were welcomed regardless of their experience with CGM. Participants were not identifiable with the data collected.

### Data collection & Survey invitation

The participant group was invited twice via the NNUH newsletter and an advertisement/invitation poster of the study was also distributed in all wards. A diabetes specialist nurse in the same hospital (DH) facilitated the distribution of the posters on the wards. Both the newsletter and the poster included a link and a QR code directing the person to the participant information sheet, consent form and the survey. We used University of East Angia (UEA) protected Microsoft Office Forms (survey) to collect the data. Our survey was open to participants for 5 months between August 2025 and December 2025. All data was anonymised and we did not collect any personal data.

### Instrument

A 38-item survey was developed to evaluate inpatient nurses’ perception and experiences of CGM, by adapted an existing questionnaire developed for ICU [16] to our potential participants from all hospital units (Appendix).

### Statistical analysis

Demographic and CGM practice survey data were analysed using RStudio (R version 4.3.2; R Foundation for Statistical Computing, Vienna, Austria). Given the nature of the study, analyses were primarily descriptive and exploratory. Descriptive statistics were used to summarise participants characteristics, personal opinions on CGM and POCT, safety and reliability of CGM and to conduct a training need analysis.

For each survey item, percentages were calculated based on the number of responses to that item. ‘N/A’ answers were excluded from the percentage calculations. Participants without previous CGM experience were not shown questions relating to CGM experience and were directed to subsequent questions using skip logic.

### Sample size considerations

Given the data collection method in our study (through invitation posters displayed across inpatient wards and newsletter circulated twice), the final sample comprised nurses who voluntarily responded to the invitation and consented to participate within the recruitment period.

### Ethics

Our project is approved by Health Research Authority (HRA) and Health and Care Research Wales (HCRW) (IRAS: 359596) in August 2025.

### Funding

We did not receive any funding for this study.

## Results

Twenty-three participants were consented during the study period. We excluded four participants since they were healthcare assistant, not nurses. Overall, nineteen in-patient nurses are included in our analysis. The average time of our online survey was approximately 5 minutes.

### Characteristics of the Participants

The majority of our participants were female (n=14, 73.7%). The other characteristics of our population is described in Table-1.

**Table-1:** Participant Characteristics.

| Characteristics | The number of participants |
| --- | --- |
| **Age (years)** |  |
| <20 | 1 |
| 20-29 | 6 |
| 30-39 | 4 |
| 40-49 | 2 |
| 50-59 | 6 |
| **Degree** |  |
| Registered nurse (RN) | 10 |
| Registered general nurse (RGN) | 3 |
| Student nurse | 3 |
| Master's degree in nursing (MPH, MHA, MBA) | 1 |
| Other* | 2 |
| **Employment** |  |
| Newly qualified nurse | 6 |
| Nursing specialist/ Senior nurse | 4 |
| Student nurse | 3 |
| Diabetes inpatient specialist nurse (DISN) | 2 |
| Advanced nurse/ nurse practitioner | 1 |
| Other** | 3 |
| **Experience in nursing practice** |  |
| Less than a year | 2 |
| 1-3 years | 2 |
| 3-5 years | 1 |
| 5-10 years | 4 |
| 10-20 years | 3 |
| More than 20 years | 4 |
| Other, student nurses | 3 |

\* Participants answered this section as 'paediatric nurse' and 'nursing associate'.
\*\* Participants answered this section as 'staff nurse', 'clinical educator', and 'nursing associate'.

### Point-of-Care Testing Experience

All participants (100%) had experience with point-of-care testing for glucose monitoring in hospital. Seventeen (89.4%) participants answered that using POCT is easy, and fourteen (73.6%) participants found it easy to document POCT readings in the electronic medical record. Fifteen (78.9%) participants found POCT reliable.

### Continuous Glucose Monitoring Experience

Eleven participants (57.9%) were familiar with CGM; ten had previously cared for people using CGM. The rest of the questions were only relevant and answered by those who had previous experience with CGM, until Question 36 which assessed all participants’ perceived need for further education on CGM. Four out of 10 (40%), reported that they cared for a patient with CGM for more than 10 shifts, five (50%) reported 1-6 shifts, and one (10%) reported they had 7-10 shifts with a patient using CGM.

### Management and Feasibility

Five participants out of seven (71.4%) agreed inserting a CGM sensor is easy and seven of nine (77.8%) found it easy to document CGM readings in the medical record. The other participants replied ‘neither agree nor disagree’ to these questions and we excluded ‘N/A’ answers to these questions. Six (66.7%) reported that CGM has helped nurses on their unit manage glucose better, while one participant (11.1%) disagreed. Six participants out of nine (66.7%) reported CGM shows novel glucose patterns that they did not see before. CGM helped to prevent any dysglycaemic event according to 66.7% of our participants (6/9). The perception on workflow followed a different pattern with only 40% of the participants (4/10) agreeing that CGM improves workflow in the hospital, two (20%) disagreed and others were neutral (4/10). Four out of seven (57.1%) found it easy to remove or replace the sensor, while others (3/7) neither agreed nor disagreed.

### Patient Care and Clinical Outcomes

Participants overwhelmingly agreed that CGM improved their patient’s care and helped them be aware of potential hypoglycaemia in advance (Chart 1). Six out of nine nurses (66.7%) reported that they prefer using a hybrid of CGM with POCT, over POCT alone.

**Chart 1.**
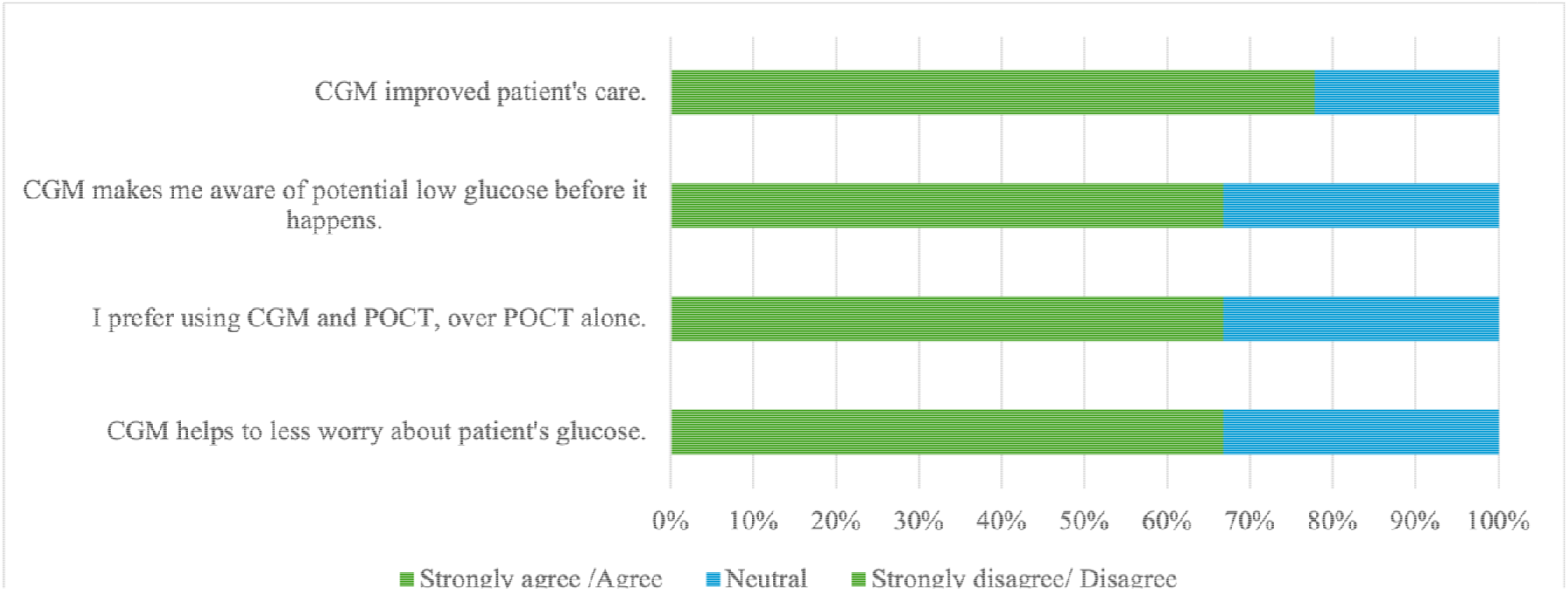
Perception on using CGM in practice based on responses from nine nurses with experience of CGM use

### Personal Opinion

Most participants found CGM reliable and accurate (7/9, 77.8%) (Chart 2). Eight out of ten (80%) reported that they are happy to use CGM in the hospital setting. Five participants out of nine (55.6%) agreed that CGM provided timely alerts and showed glucose levels to be predictable. However, there was an equivalence between nurses who agreed that CGM reduces the time of entries to patient’s room (n=3, 33.3%) and those who disagreed with this statement (n=3, 33.3%). Nearly all nurses (n=9, 90%) reported that they would recommend using CGM to other colleagues.

**Chart 2.**
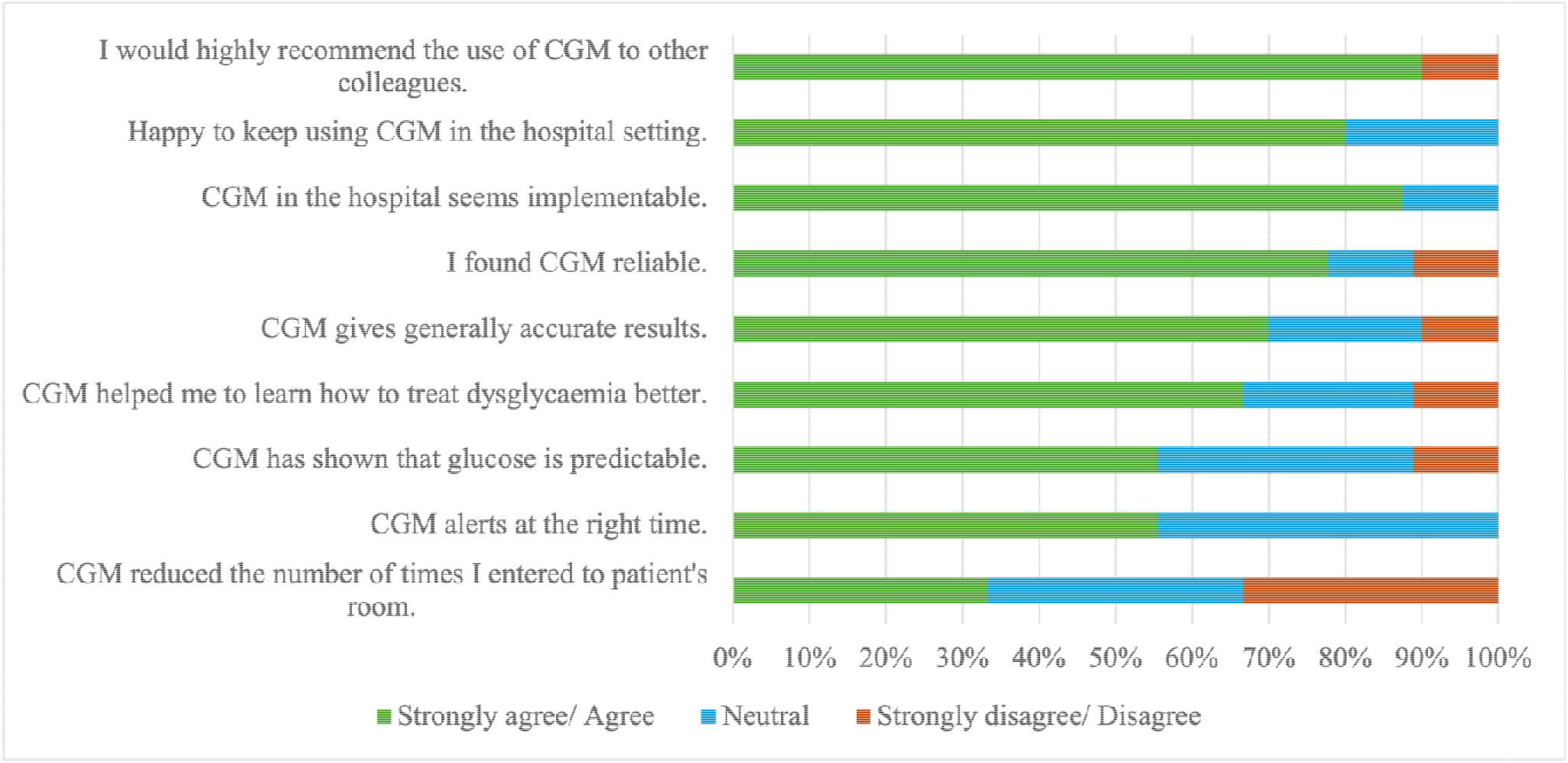
Nurses’ personal opinions and their perceptions on accuracy and reliability of CGM

### Training Assessment of Nurses and Training Need Analysis

CGM learning resources consisted of training from another nurse, online training videos and other training materials by suppliers. Eleven nurses answered this multiple-option question. In nine of eighteen responses (50%), nurses had training from either another staff nurse familiar with CGM (5 answers), a charge nurse (1) answer) or a diabetes specialist nurse (3 answers). Three responses (16.7%) indicated that nurses learned from online training videos, two (11.1%) from reading the protocol, and one (5.6%) from product suppliers. Two responses (11.1%) showed that nurses had CGM training as part of their annual proficiency training. One participant (5.6%) learned from a parent caring for their child living with diabetes.

Ten (90.9% of 11) nurses reported their training on CGM was sufficient, however, thirteen (68.4%) of total participants (n=19) wanted to become familiar with CGM and reported they would like to have more information about sensors.

All participants (100%) answered training need analysis questions. Participants were allowed to select multiple options. The results showed that nurses most need (68.4% of participants, 28.3% of all answers) further training on inserting and replacing (placement) of CGM, which comparable to the training need of understanding alarms and alerts of sensors (57.8% of participants). Participants also needed to learn more about the scientific background and reliability of CGM (52.6%), and only four participants (21%) reported that they did not need further training. We summarised the training need analysis results in Chart 4.

**Chart 3.**
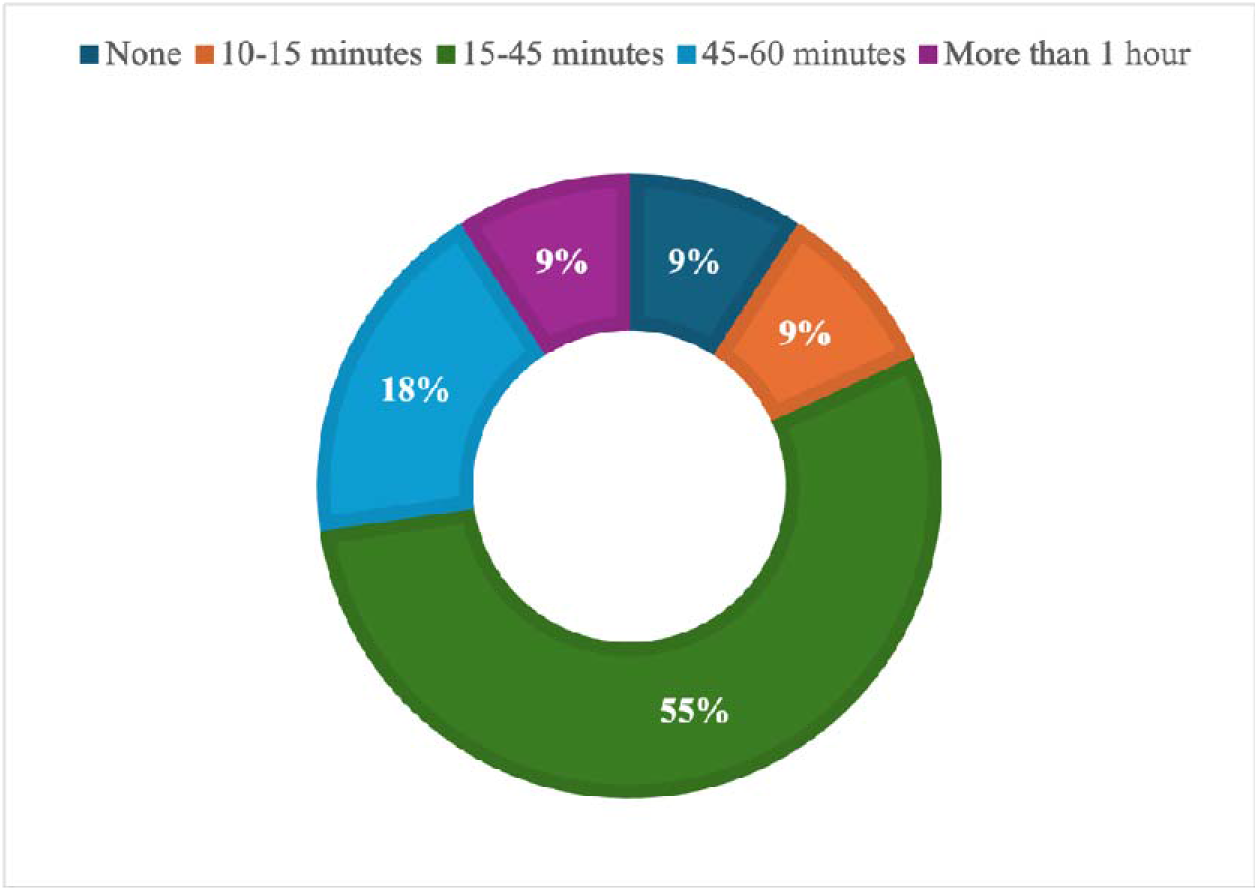
Time spent on CGM training

**Chart 4.**
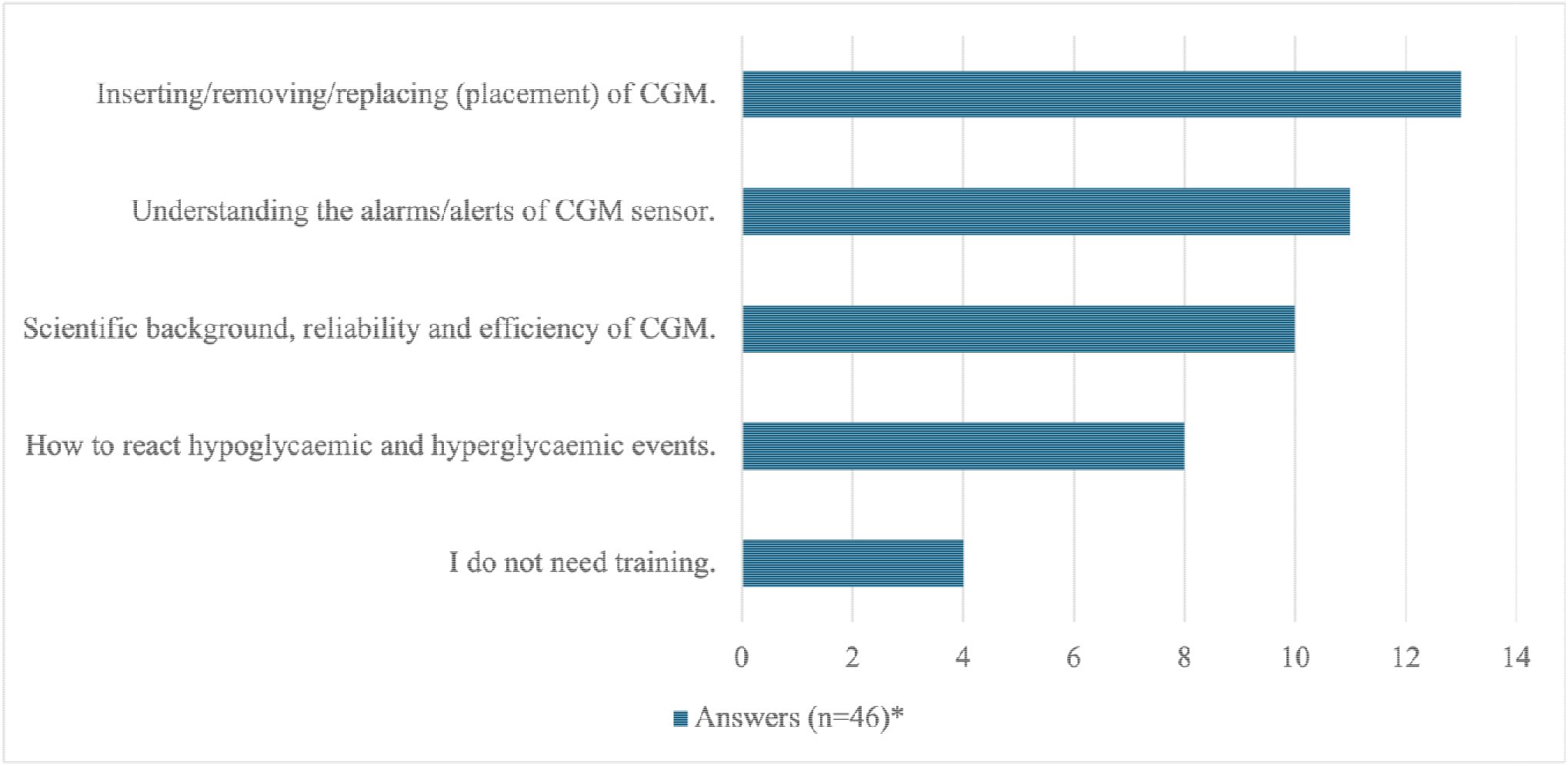
Self-assessment of nurses on CGM training need analysis *Participants (n=19) were allowed to select multiple options. The total number of answers were 46.

Lastly, participants were invited to provide additional comments or suggestions through an open-ended question at the end of the online survey. Free-text responses highlighted both the perceived clinical value of CGM use in hospital settings as well as some concerns regarding its reliability compared to POCT. There were three answers to this question.

*“I believe CGM will become more prevalent in future DM monitoring therefore to have the underpinning knowledge in their use will be beneficial in providing relevant nursing care.”* – ID1.

*“From personal experience, the CGM’s are showing different readings compared to the trust glucose machines.”* – ID3.

*“We are advised to use the Trust POCT machines that have been calibrated rather than allowing patients who come into hospital with CGM’s to tell us what their blood sugars are (paediatrics). This, in my opinion, is a waste of resources.”* – ID8.

## Discussion

Our findings provide a first insight into how general nursing staff experience and perceive CGM and POCT in hospital settings. More than half of the nurses had a caring experience with CGM within their inpatient practice. The majority of the nurses found it easy to insert a CGM sensor and document the readings. CGM was considered a reliable and accurate tool which allowed the nursing team to be aware of potential hypoglycaemic events in advance. Most of the nurses preferred using the CGM + POCT combination, rather than POCT alone. However, nurses expressed mixed views regarding whether CGM improved the workload in the hospital or reduced the frequency of patient room entries, suggesting uncertainty about its impact on workload. In terms of training needs, nurses needed to learn more on sensor placement and how to act upon or triage hypo-or hyperglycaemic alerts of CGM sensors.

Overall, the nurses who had experience with CGM were very positive towards the technology, even without having a specialist background in diabetes. In addition, most nurses agreed that inserting CGM is easy, however, at the same time, the majority of respondents also reported further training needs for sensor placement. This finding suggests that although CGM insertion is generally perceived as easy, structured training remains essential to ensure consistent and accurate sensor placement, as suggested in the previous reports highlighting the importance of education in CGM implementation [19]. We also replicated previous findings in ICU nurses [16], showing that most nurses prefer the use of CGM alongside POCT, rather than CGM or POCT alone. The reason for this is not clear, although it seems there are concerns about reliability of CGM readings compared to POCT. This is despite extensive reliability studies evidence showing acceptable agreement between CGM and POCT [20], [21]. Further training on reliability findings might alleviate the concerns of CGM reliability in nurses, although POCT readings should still be obtained in the event of hypoglycaemic events, as recommended by most healthcare service guidelines. Still, CGM could have clear advantages compared to current POCT standard of care, not only in reduced patient discomfort, due to repeated finger pricking but also in the closer monitoring of highly vulnerable populations, which are common in hospital settings [19].

In particular the detection of dysglycaemic events might help better clinical management of vulnerable inpatients. The majority of nurses within our survey with CGM experience recognised this advantage and reported that CGM increases their awareness of low glucose readings before it occurs. Hypoglycaemia (<3.9 mmol/L) is the most concerning short-term complication for people living with diabetes, carers and healthcare professionals [22]. Therefore, there are a number of CGM implementation studies conducted in high-risk population for hypoglycaemia, showing hypoglycaemia can be reduced with the use of CGM [19], [23], [24], [25]. Previous research on nursing practice demonstrated that CGM helped them to worry less about patient’s potential hypoglycaemia due to early detection of CGM. Nurses reported that CGM is feasible and efficient in the hospital setting [26]. Our findings replicate that and show that even general nursing staff are aware of the potential benefits of CGM to monitor glycaemic levels with increased beneficial outcomes for inpatients.

However, at the same time, the nurses in our survey expressed mixed opinions whether CGM decreases their workload and the entries to patient room. In our findings, the percentage of nurses indicating that CGM reduce the entering of rooms showed an equal distribution between agree, no change and disagree. It might depend on the ward or patient characteristics whether CGM decreased the workloads. This would chime with the ICU findings, where it clearly reduced attendance, likely due to higher intensity of glucose monitoring required in intensive care [16]. However, in general wards, where glucose checks are only recommended, before meals and at bedtime, which may explain this inconsistent observation [24], [27]. These findings raise the question, as to whether guidance on CGM alerts is required for general wards, since they allow earlier detection of hypoglycaemia while potentially increasing the attendance frequency of nurses. Future research is clearly needed to determine whether CGM provides better overall clinical outcomes and consequently mitigates the potential increase in healthcare attendance across different wards or patient populations.

In terms of reported barriers for CGM use in hospital settings, previous studies demonstrated high consumable costs of sensors and insufficient insurance cover and lack of sufficient training for healthcare professionals [28]. In terms of training, our findings showed that the majority of nurses in our survey reported to have had sufficient training for CGM (90.9%) which is similar to previous research with ICU nurses (80%) [16]. However, at the same time, over 68% of the nurses also expressed they would like to have more information on CGM and become more familiar with its use, in particular for clinical decision-making, replicating previous findings [29].

Therefore, detailed training need analysis plays essential role to assess the need and develop training to help wide use of CGM in both hospital and community settings. Our findings suggest that inpatient general nursing staff had less confidence in sensor placement compared to other training need areas and they reported that further training on inserting, replacing or removing CGM is needed. Another survey with DSNs showed a lack of structured NHS-based programmes for CGM-integrated hybrid closed-loop (HCL) systems [17]. DSNs highlighted a lack of hands-on training with for HCLs, with learning largely dependent on industry-led sessions rather than structured practical training [17]. Our general nursing findings replicate these findings in diabetes specialist nursing and highlight that general nurses might require further training specifically tailored for their needs. Along the same lines, reported safety concerns and confusion regarding alarms/alerts of CGM among general healthcare professionals highlight the need for further training [17], [30]. Overall, these results suggest that insufficient practical experience and training among healthcare professionals may represent a barrier to CGM adoption in hospital settings.

Despite our interesting and novel findings, our study had several limitations. First, the study was constrained to a single tertiary hospital with a relatively small sample size reflecting the number of nurses who voluntarily participated during the recruitment period. This may limit the generalisability of the findings and highlights the need for a replication study in multiple centres and with larger general nurse populations. Still, our quick survey allows for rapid scaling up and implementing in various settings, which should overcome this limitation in the near future. Further, the surveys captured self-reported perceptions rather than objective measures of CGM use, workload or clinical outcomes, which might have led to response biases, common in survey studies. Still, the survey allowed nurses to contribute to the research even without being involved in research studies. Having the voice of the general nursing staff represented is important as they come across CGM sensors in a regular basis in their clinical duty, even without objective confirmation of CGM usage. Lastly, nurses were recruited on a voluntary basis, which may have caused a self-selection bias. Specifically, nurses with a greater interest in diabetes technology and prior exposure to CGM use may have been more likely to participate. Such selection bias is, unfortunately, common in voluntary research. However, some of our findings are consistent with previous other studies, suggesting that similar findings can be obtained from different populations, which may mitigate concerns about selection bias.

Overall, inpatient nurses expressed largely favourable perceptions on CGM and preferred its use alongside POCT for glucose monitoring. While the impact of CGM on workload remains uncertain, addressing training needs on sensor placement and alarm management may facilitate successful implementation in inpatient care. Multicentre studies with larger and diverse nursing populations are needed to confirm these findings and identify the training and organisational requirements needed for the wider adoption of CGM in hospital settings.

## Data Availability

All data produced in the present study are available upon reasonable request to the authors.

